# Long-term symptoms after SARS-CoV-2 infection in a cohort of hospital employees: duration and predictive factors

**DOI:** 10.1101/2022.03.22.22272770

**Authors:** Rosalie Gruber, Maria Veronica Montilva Ludewig, Christina Weßels, Gerlinde Schlang, Svenja Jedhoff, Swetlana Herbrandt, Frauke Mattner

**Author notes:** <u>Corresponding author:</u> Rosalie Gruber, Institute of Hygiene, Cologne Merheim Medical Center, University Hospital of Witten/Herdecke, Ostmerheimerstr. 200, 51109 Köln, Germany.

## Abstract

**Objective:** To evaluate the frequency, duration and patterns of long-term coronavirus disease 2019 (COVID-19) symptoms and to analyze risk factors for long-lasting COVID-19 sequelae among hospital employees (HEs).

**Design:** Retrospective observational cohort study.

**Setting:** Three medical centers in Cologne, Germany.

**Participants:** We included HEs who had a positive severe acute respiratory syndrome coronavirus 2 (SARS-CoV-2) polymerase chain reaction (PCR) test between March 2020 and May 2021.

**Methods:** We conducted a survey in mid-2021 with all HEs tested SARS-CoV-2 PCR positive during the study period and asked about the presence and duration of 24 different COVID-19 related symptoms. Chronological development and patterns of symptom complexes, probability of symptom persistence and possible risk factors for protracted COVID-19 course were analyzed.

**Results:** Of 221 included HEs, a number of 104 HEs (47.1%) reported at least one persisting symptom for more than 90 days after initial SARS-CoV-2 detection. A symptom duration over 28 days was associated with multiple symptom complexes. The most common was the interrelated occurrence of shortness of breath, memory disorder, concentration disorders and fatigue. Each one cycle higher initial Ct value significantly increased the chances of overcoming symptoms (odds ratio [OR] = 1.05; p = 0.019). The occurrence of breathlessness within the first ten days (OR = 7.89; p = 0.008), an initial Ct value under 30 (OR = 3.36; p = 0.022) and a definitely nosocomial SARS-CoV-2 transmission (OR = 3.05; p = 0.049) showed a statistically significant association with increased odds of illness duration over 90 days.

**Conclusion:** About half of the HEs suffered from long lasting symptoms over 90 days after almost entirely mild acute COVID-19. Different symptom complexes could be shown and predictive factors for long-term symptoms were identified. Predictive factors at the onset of the infection could possibly be used for early treatment to prevent development of long-term symptoms after COVID-19 in future.

## Introduction

After the third wave of severe acute respiratory syndrome coronavirus type 2 (SARS-CoV-2) infections in springtime 2021 in Germany, we observed at University Hospital Cologne-Merheim a high number of infected hospital employees (HE) with long-lasting symptoms after predominantly mild acute courses of coronavirus disease 2019 (COVID-19).^1,2^ In contrast to more frequently studied long-lasting symptoms after severe COVID-19 courses, ^3-5^ we were surprised by the amount of symptoms that also occurred after mild courses not requiring hospitalization. Since the vast majority of people infected with SARS-CoV-2 do not need to be hospitalized,^6,7^ it is of great importance to also consider mild COVID-19 cases with regard to long-lasting COVID-19 symptoms.

There are numerous different terms and definitions for long-lasting COVID-19 symptoms. The World Health Organization developed the following clinical case definition: “Post COVID- 19 condition occurs in individuals with a history of probable or confirmed SARS-CoV-2 infection, usually 3 months from the onset of COVID-19 with symptoms that last for at least 2 months and cannot be explained by an alternative diagnosis”.^8^

Here, we had the opportunity to investigate the entire cohort of SARS-CoV-2 infected HEs. HEs comprise a quasi-representative cohort of the employable population, here in Cologne in Germany. Furthermore, SARS-CoV-2 positive HEs were not only detected by testing due to respiratory or influenza-like symptoms but also by active screening of all suspected contacts to known SARS-CoV-2 cases in either patients, colleagues, friends, or family members. Therefore, we reasoned that the cohort would be appropriate to study the variety, combination, and duration of symptoms after non-severe courses of COVID-19. Hence, we started standardized phone interviews in June 2021 with all HEs who have had a SARS-CoV-2 positive PCR test since the beginning of the pandemic. We asked about the presence of different symptoms and their respective durations and linked the information to data of the index and contact management available in the Institute of Hygiene. Thus, a description of the combination of different symptoms over time and risk factor analysis for a prolonged course after non-severe COVID-19 was enabled.

## Methods

### Study setting

A retrospective observational cohort study was conducted among SARS-CoV-2 positive hospital employees (HEs) of three medical centers in Cologne, Germany.

### Data collection

All HEs who had a positive SARS-CoV-2 PCR test from the beginning of the pandemic in March 2020 until May 2021 were recorded by staff of the hospitals’ Institute of Hygiene. The following data was collected for each HE after positive SARS-CoV-2 PCR detection: name, gender, date of birth, address, phone number and/or E-mail address, date of SARS-CoV-2 detection, Ct values of multiple PCR testings, initial symptoms, contact persons and contact situations, vaccination status, data on the inpatient course in in the event of hospital admission. This data was required for fulfilment of the reporting obligations to the city’s public health department. From the beginning of the pandemic, the Occupational Health Service of the three medical centers looked after HEs who had been infected with SARS-CoV-2 and in some cases suffered from long-lasting COVID-19 symptoms. All HEs with previous SARS-CoV-2 infection were contacted by phone and asked about their willingness to participate in a systematic survey on possible long-term COVID-19 symptoms. HEs were given the option to cancel the survey at any time. The survey was conducted by staff of both the Occupational Health Service and the Institute of Hygiene and was carried out between June and October 2021. All HEs interviewed had at least a six-week interval from their first SARS-CoV-2 positive PCR result at the time of the interview. In preparation for the survey, a structured questionnaire was jointly developed that included 24 different long-term COVID-19 symptoms that were frequently described in previously published studies about long-lasting COVID-19 symptoms,^7,9,10^and adapted to individual symptoms of the HEs (supplemental material). HEs were asked only about symptoms that occurred after SARS-CoV-2 detection and were not asked to report symptoms that had previously existed. The self-assessed symptom severity (mild/medium/severe) and the respective time of symptom onset and end (beginning/middle/end of a month) were asked. In addition, three different sources of infection were defined by physicians of the Institute of Hygiene and applied to each HE’s case. A definite nosocomial infection of HEs with SARS-CoV-2 was, by definition, present when there was a defined outbreak on a medical ward with exactly fitting time period. Contact with a COVID-19 case within the hospital with missing or inappropriate personal protective equipment within the last 14 days was also classified as a definite nosocomial infection. In the event of contact with a COVID-19 case within the hospital with appropriate personal protective equipment and lack of private contact to a COVID-19 case within the last 14 days, the infection was categorized as possible nosocomial infection. If there was contact with a COVID-19 case in the private setting or there was no contact situation within the healthcare facility within the last 14 days, this was categorized as a community-acquired COVID-19 infection.

### Data analysis

Descriptive analysis of frequency and duration of long-term COVID-19 symptoms were conducted. Analysis was performed using the Statistical Software R.^11^ Using co-occurrence network analysis, the formation, patterns and chronology of COVID-19 symptom complexes after SARS-CoV-2 infection were assessed.^12^ Survival-time analysis was conducted in order to assess the probability of respective symptom persistence using Kaplan-Meier estimator.^13^ Likewise, univariate analysis of symptom durations depending on professional group, age, initial Ct value and other influencing factors was carried out using Cox regression.^14^ Possible risk factors for a protracted COVID-19 course with long-lasting COVID-19 symptoms were identified using logistic regression.^15^ P values <.05 were considered statistically significant.

### Ethical clearance

This study was approved by the ethics committee of the Witten/Herdecke University (S-273/2021). The study was core funded.

## Results

In total, 297 HEs tested positive for SARS-CoV-2 during our study period of which 221 HEs were interviewed for the survey. A number of 76 HEs were excluded because of survey refusal, survey drop out or incomplete survey. Ct values were available for 150 HEs (67.9%) (Table 1). Mean age was 39.76 years (standard deviation (SD), 12.46; age range 17-64 years) and 158 HEs (71.5%) were female. During the acute COVID-19 infection, five HEs (2.3%) received inpatient care, including two HEs (0.9%) who received intensive care treatment. At the time of SARS-CoV-2 positivity, seven HEs (3.2%) had been vaccinated twice against COVID-19.

**Table 1.** Characteristics Of Investigated Cohort Of SARS-CoV-2 Positive Hospital Employees.

|  | Frequency |  | Symptom Duration, days |  |  |
| --- | --- | --- | --- | --- | --- |
|  | N | % | Median (IQR) | Mean | Min-Max |
| Total number | 221 | 100.0 | 112 (210.0) | 139.9 | 0-491 |
| **Demographics** |  |  |  |  |  |
| Female | 158 | 71.5 | 129 (209.8) | 146.5 | 0-491 |
| **Age** |  |  |  |  |  |
| 16-30 | 70 | 31.7 | 70 (221.3) | 116.6 | 0-474 |
| 31-40 | 44 | 19.9 | 83.5 (184.5) | 144.3 | 0-482 |
| 41-50 | 55 | 24.9 | 145 (202.0) | 137.0 | 0-491 |
| 51-65 | 52 | 23.5 | 168 (175.5) | 170.4 | 0-482 |
| **Professional group** |  |  |  |  |  |
| Nursing staff | 117 | 52.9 | 147 (214.0) | 149.7 | 0-491 |
| Medical staff | 24 | 10.9 | 50 (197.5) | 128.5 | 0-462 |
| Others (e.g., administration, cleaning staff) | 80 | 36.2 | 87.5 (194.3) | 128.9 | 0-474 |
| **Working on a COVID ward** | 59 | 27.0 | 182 (200.5) | 162.5 | 0-471 |
| **Transmission type** |  |  |  |  |  |
| Community acquired | 114 | 51.6 | 57.5 (198.5) | 117.5 | 0-491 |
| Definitely nosocomial | 62 | 28.0 | 177 (168.5) | 165.8 | 0-482 |
| Possibly nosocomial | 45 | 20.4 | 183 (180.0) | 160.8 | 0-471 |
| **Initial Ct value available** | 150 | 67.9 |  |  |  |
| **Duration of COVID symptoms** |  |  |  |  |  |
| No persisting symptoms at the date of the interview | 111 | 50.2 | 22.0 | 47.4 | 0-397 |
| At least one persisting symptom at the date of the interview | 110 | 49.8 | 224.5 | 233.2 | 58-491 |

Note. IQR, interquartile range. Study population: hospital employees of three medical centers in Cologne, Germany, with SARS-CoV-2 infection between March 2020 and May 2021.

After SARS-CoV-2 detection, 27 HEs (12.2%) reported no symptoms at all. In 33 HEs (14.9%), at least one symptom occurred for a minimum of one day and lasted no longer than 28 days. More than 90 days after initial SARS-CoV-2 detection, 104 HEs (47.1%) reported at least one persisting symptom. The most common symptoms were fatigue (n = 139, 62.9%), headache (n = 121, 54.8%), anosmia (n = 120, 54.3%) and ageusia (n = 118, 53.4%) (Table 2). With a median symptom duration of six days (mean = 7.7 days; SD = 8.49), fever represented the symptom with the shortest median symptom duration. The symptoms with the longest median symptom duration were alopecia with a median of 158.5 days (mean = 171.8 days; SD = 118.96) and memory disorder with a median of 155 days (mean = 158.5 days; SD = 112.73).

**Table 2:** Symptom Frequency And Duration After SARS-CoV-2 Positivity.

| **Symptoms** | **Frequency**<br>**(N=221)** | **Symptom Duration (days)** |  |  |
| --- | --- | --- | --- | --- |
|  | **N (%)** | **Mean** | **Median** | **SD** |
| Fever | 81 (36.7) | 7.7 | 6.0 | 8.49 |
| Rhinitis | 62 (28.1) | 31.2 | 10.5 | 76.35 |
| Sore throat | 78 (35.3) | 38.9 | 8.0 | 91.04 |
| Limb pain | 70 (31.7) | 27.6 | 9.0 | 49.85 |
| Cough | 89 (40.3) | 46.0 | 16.0 | 84.02 |
| Weakness in the limbs | 78 (35.3) | 66.1 | 15.0 | 100.80 |
| Heaviness of limbs | 67 (30.3) | 53.2 | 15.0 | 74.89 |
| Headache | 121 (54.8) | 62.1 | 15.0 | 104.08 |
| Brain fog | 54 (24.4) | 71.9 | 15.0 | 115.57 |
| Skin alterations | 17 (7.7) | 66.2 | 25.0 | 84.78 |
| Anxieties | 43 (19.5) | 89.0 | 26.0 | 101.37 |
| Dysesthesia | 25 (11.3) | 105.0 | 32.0 | 129.64 |
| Fatigue | 139 (62.9) | 95.8 | 36.0 | 114.83 |
| Ageusia | 118 (53.4) | 81.1 | 32.0 | 105.19 |
| Anosmia | 120 (54.3) | 85.8 | 35.0 | 103.96 |
| Unspecific pain | 29 (13.1) | 98.7 | 60.0 | 111.92 |
| Breathlessness | 50 (22.6) | 120.3 | 63.5 | 139.68 |
| Sleep disorder | 54 (24.4) | 133.6 | 83.0 | 137.48 |
| Palpitations | 57 (25.8) | 127.4 | 64.0 | 140.46 |
| Shortness of breath | 105 (47.5) | 118.4 | 83.0 | 119.08 |
| Concentration disorders | 94 (42.5) | 127.0 | 94.0 | 116.58 |
| Mood changes | 26 (11.8) | 148.0 | 140.0 | 134.12 |
| Memory disorder | 57 (25.8) | 158.5 | 155.0 | 112.73 |
| Alopecia | 24 (10.9) | 171.8 | 158.5 | 118.96 |

Note. SD, standard deviation. Study population: hospital employees of three medical centers in Cologne, Germany, with SARS-CoV-2 positivity between March 2020 and May 2021.

Using co-occurrence network analysis, the formation of symptom complexes were demonstrated (Figure 1; Figure 2; supplementary movie). For HEs who had a symptom duration of less than 28 days, the most prominent combined symptoms were ageusia and anosmia (Figure 1; Figure 3). With a symptom duration over 28 days, a formation of additional network points of symptom complexes such as fatigue or concentration disorders, among others, could be seen. The most prominent symptom complex was the interrelated occurrence of shortness of breath, memory disorder, concentration disorders and fatigue (Figure 2; Figure 4).

**Fig. 1.**
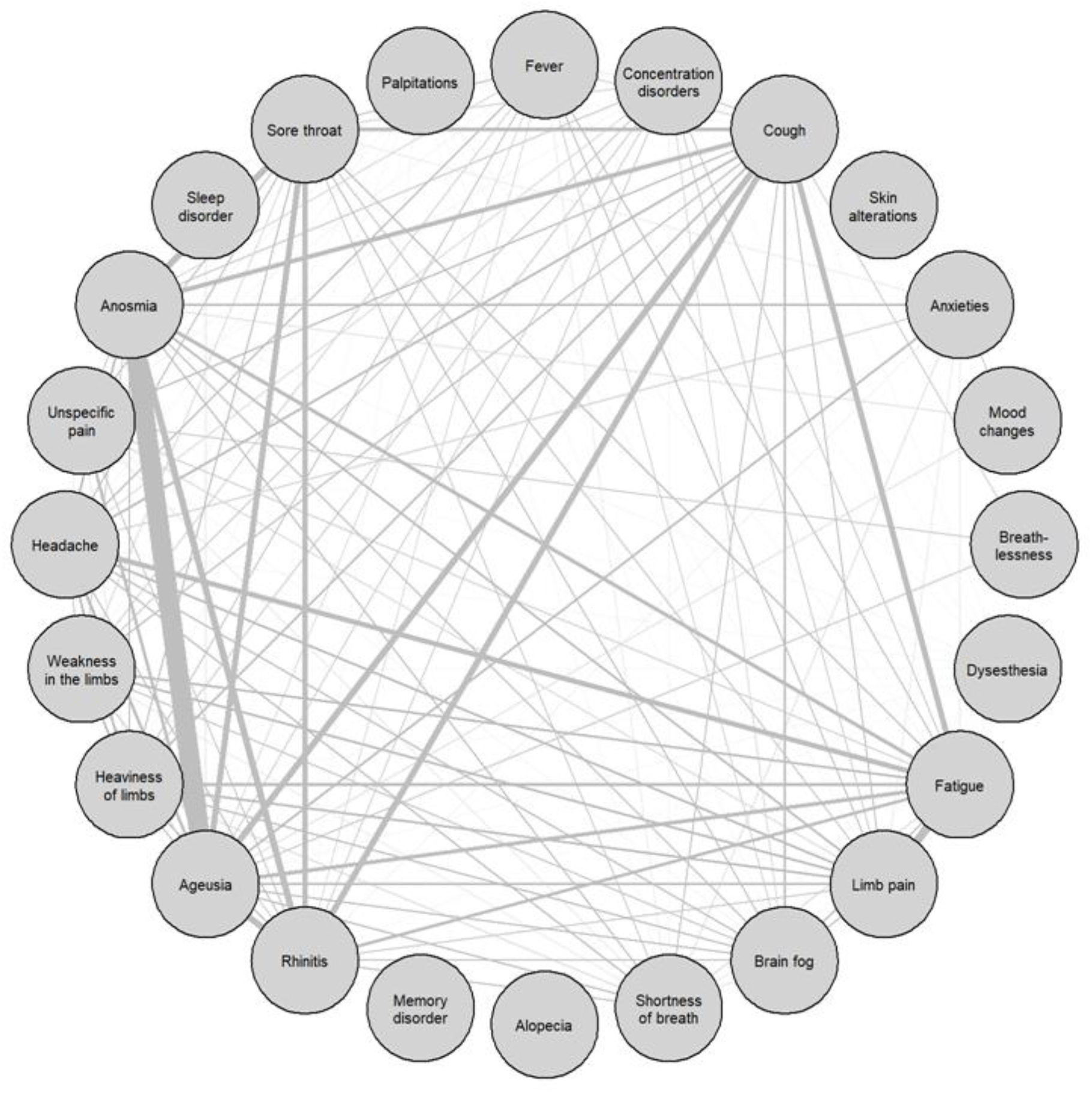
Co-occurrence network of symptoms. Symptom network includes symptoms only of hospital employees with symptom duration of max. 28 days (N=33). Each line shows the simultaneous occurrence of the respective symptoms. The thicker the line, the more often symptoms occurred simultaneously.

**Fig. 2.**
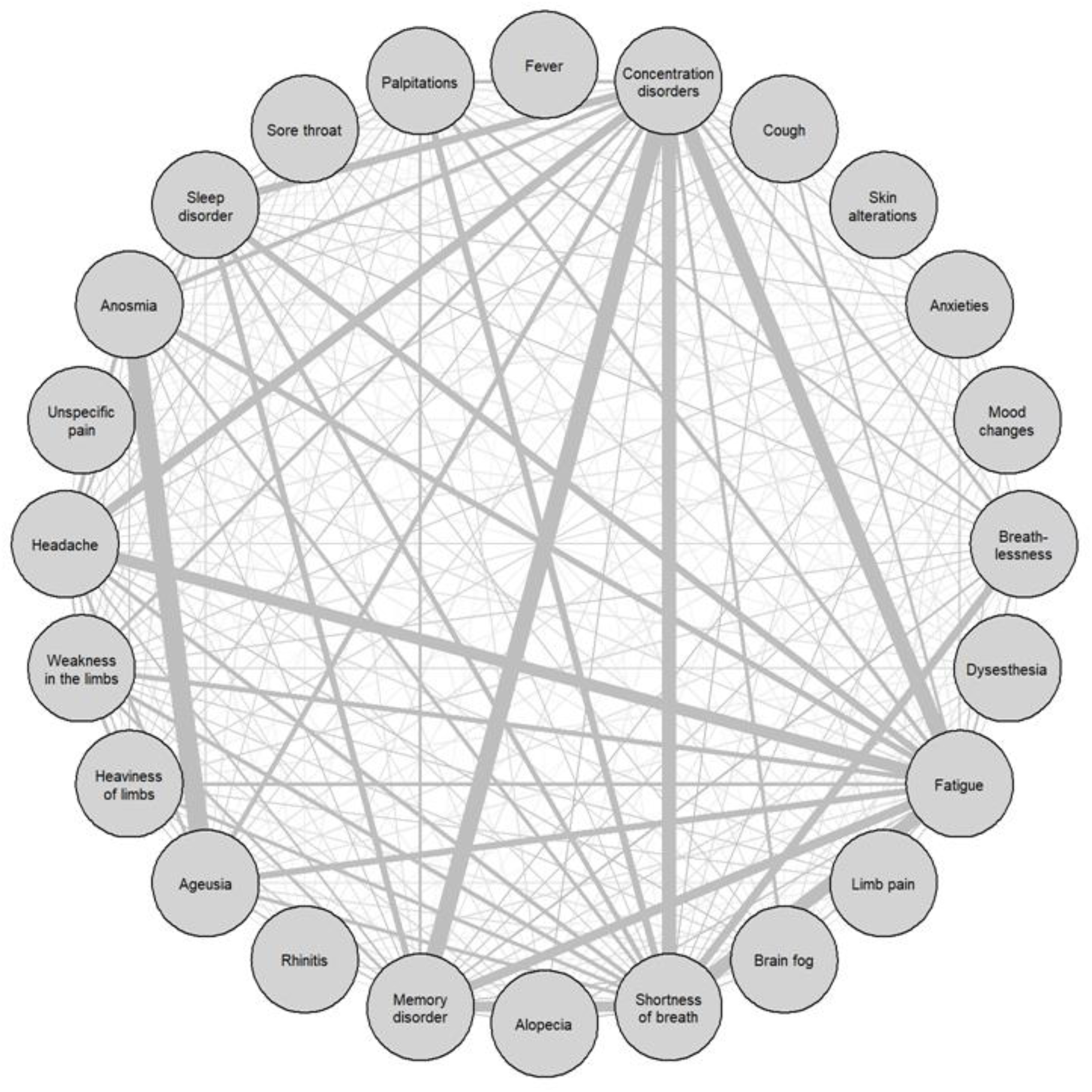
Co-occurrence network of symptoms. Symptom network includes symptoms only of hospital employees with symptom duration more than 28 days (N=161). Each line shows the simultaneous occurrence of the respective symptoms. The thicker the line, the more often symptoms occurred simultaneously.

**Fig. 3:**
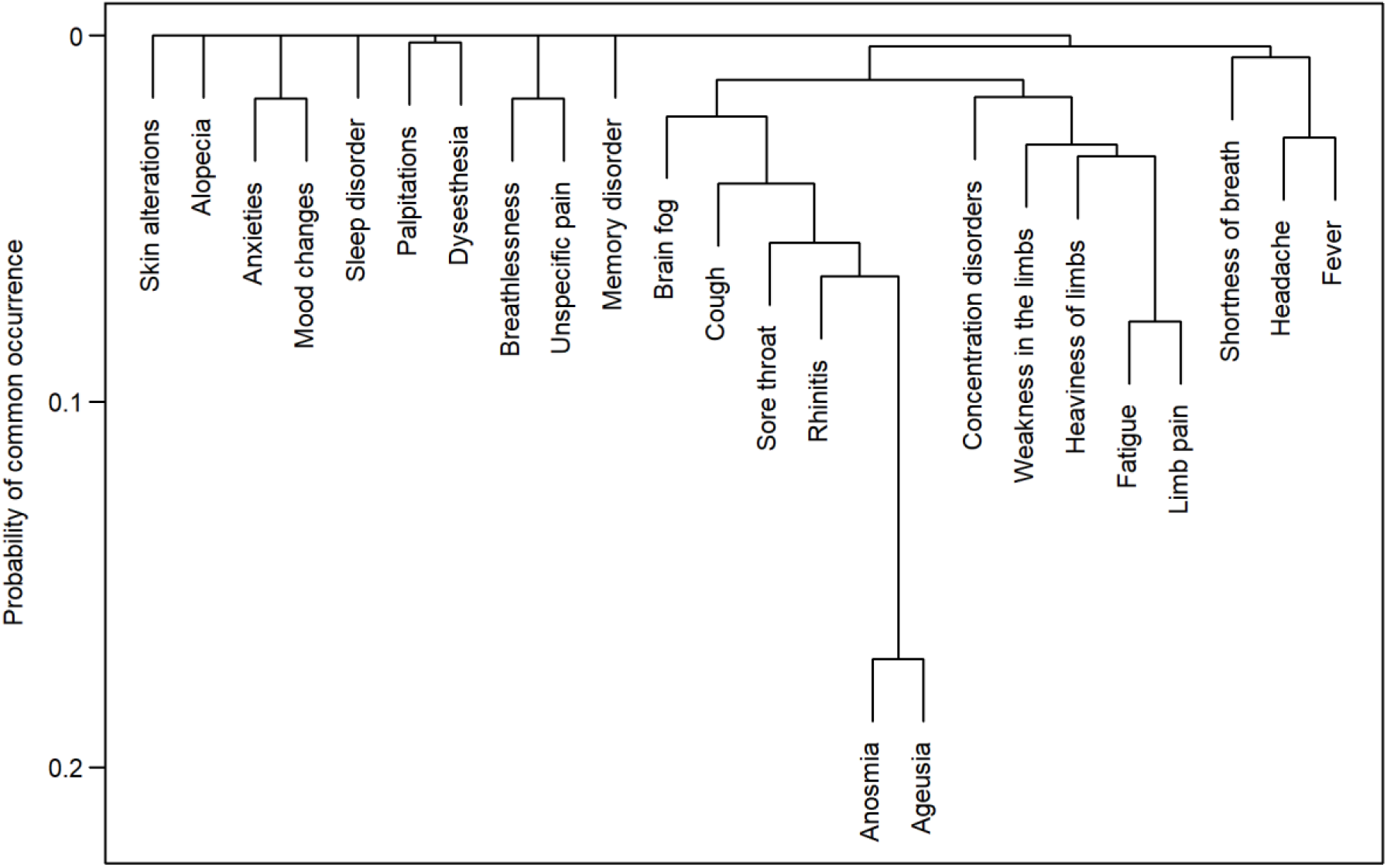
Probability of common occurrence of COVID-19 symptoms. This symptom dendrogram, which is a hierarchical clustering with complete linkage, includes only data of hospital employees with symptom duration of max. 28 days (N=33).

**Fig. 4:**
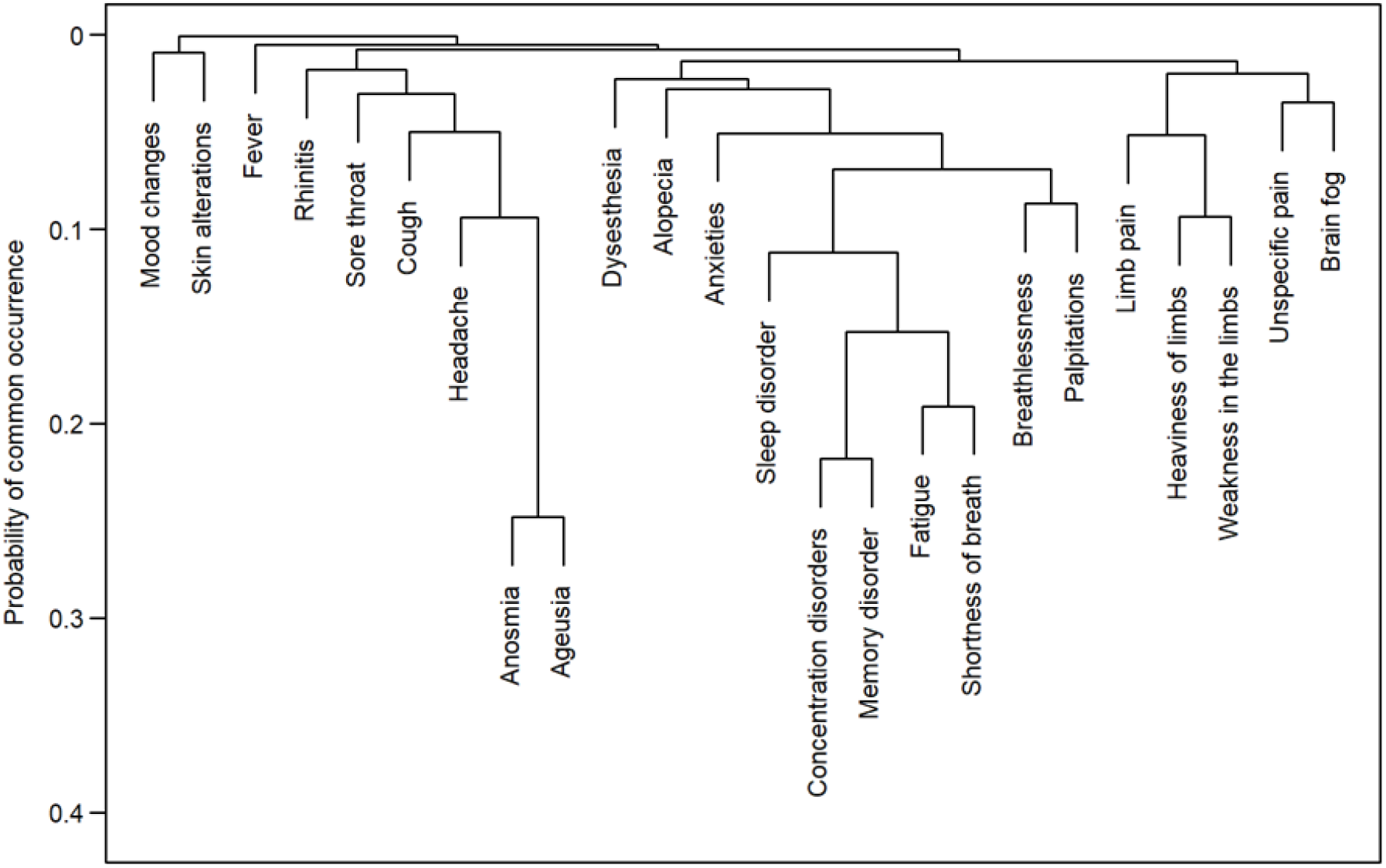
Probability of common occurrence of COVID-19 symptoms. This symptom dendrogram, which is a hierarchical clustering with complete linkage, includes only data of hospital employees with symptom duration over 28 days (N=161).

The probability of symptom persistence was plotted using Kaplan-Meier curves (Figure 5). The symptom least likely to still be present after 90 days was fever (0.0% of n = 81), whereas alopecia was the symptom most likely to still be present after 90 days (78.0% of n = 24). For HEs who developed shortness of breath after SARS-CoV-2 detection (n = 105), the probability for persisting shortness of breath after 200 days was 39.0%. For HEs suffering from memory disorder (n = 57), the probability for persistence of memory disorder after 200 days was 58.4%.

**Fig. 5.**
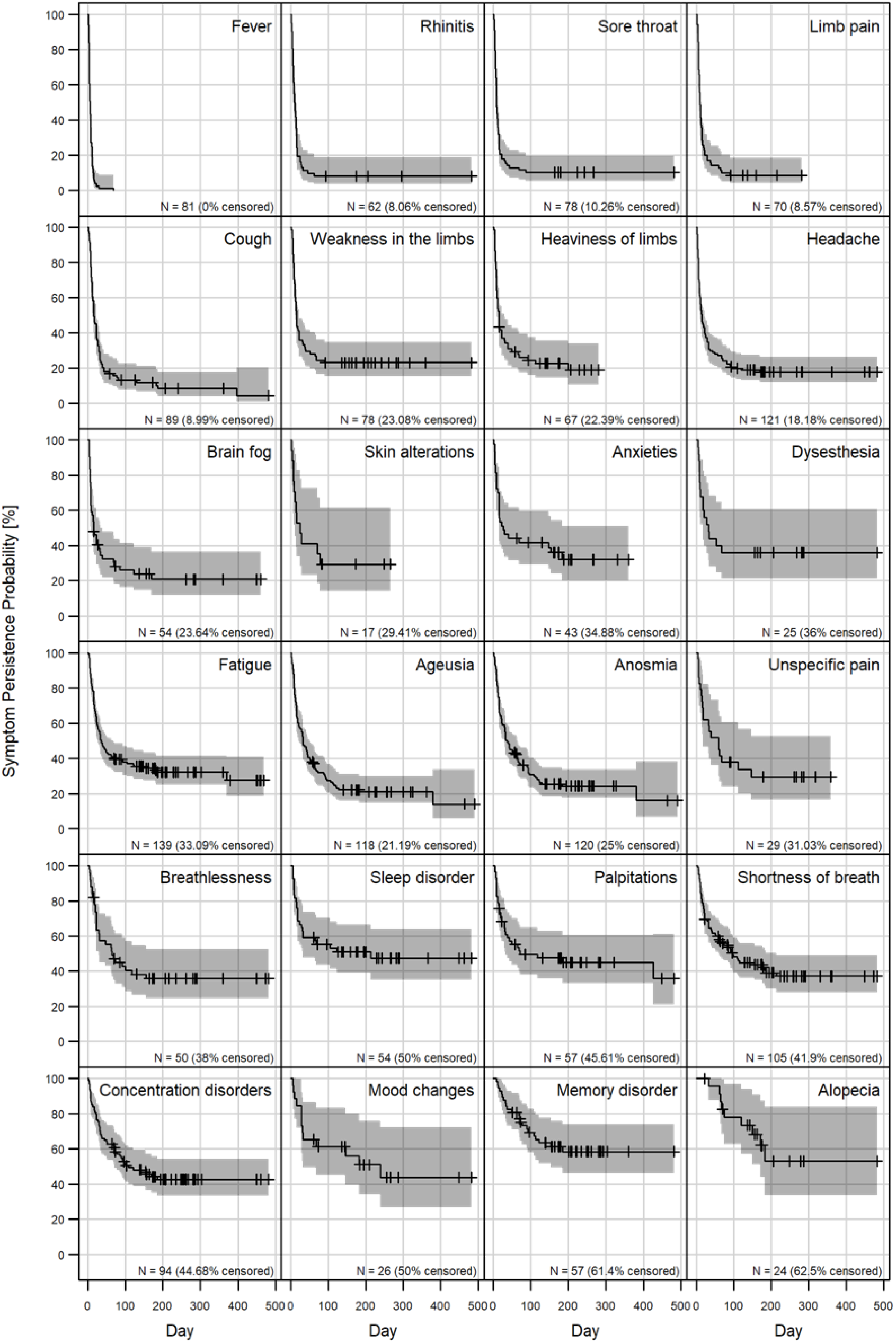
Kaplan-Meier curves of probability of symptom persistence. For each symptom, the number of hospital employees (N) who developed the respective symptom is stated. The percentage of censored data indicates how many hospital employees still had the respective symptom at the time of the survey. All graphs were arranged from top left to bottom right with increasing 28-days survival probability of the respective symptom.

Cox regression analysis showed that each one cycle higher initial Ct value significantly increased the chances of overcoming symptoms (odds ratio [OR] = 1.05; p = 0.019) (Table 3). The chances of being symptom-free for HEs who had a possibly nosocomial infection were significantly lower than for HEs with a community acquired SARS-CoV-2 infection (OR = 0.48; p = 0.044). The Cox regression model also indicated that there was a higher chance of symptom resolving for medical staff than for nursing staff (OR = 2.13; p = 0.055).

**Table 3:** Cox Regression Model For The Event Odds Of Becoming Asymptomatic.

|  | Odds Ratio | SE | p value |
| --- | --- | --- | --- |
| Gender (female) | 0.76 | 0.28 | 0.319 |
| Age at the time of COVID disease | 0.99 | 0.01 | 0.376 |
| Professional group (Medical staff) | 2.13 | 0.39 | 0.055 |
| Professional group (Others) | 1.02 | 0.30 | 0.938 |
| Work on the COVID ward (yes) | 0.89 | 0.32 | 0.711 |
| Initial Ct value | 1.05 | 0.02 | **0.019** |
| Transmission type (Possibly nosocomial) | 0.48 | 0.36 | **0.044** |
| Transmission type (Definitely nosocomial) | 0.59 | 0.30 | 0.074 |

Note. SE; standard error.
The event odds of becoming asymptomatic at day t are modeled. Only hospital employees with available initial Ct value were included (N=150). For nominal scaled variables (gender, professional group, work on the COVID ward, transmission type), odds ratio refers to the reference characteristic (male gender, nursing staff, work on the non-COVID ward, community acquired). For interval scaled variables (age, Ct value), odds ratio refers to the characteristic that is one point/cycle higher in each case.
To avoid misleading interpretation and since the modelled outcome is a positive event (asymptomatic), we refrain from hazard terminology in favor of odds.

**Table 4:** Odds For Exceeding Illness Duration Of 90 Days Based On Logistic Regression.

|  | Odds Ratio | SE | p value |
| --- | --- | --- | --- |
| Age (31-40) | 0.97 | 0.64 | 0.960 |
| Age (41-50) | 1.96 | 0.59 | 0.256 |
| Age (51-65) | 3.03 | 0.65 | 0.089 |
| Gender (female) | 1.99 | 0.55 | 0.212 |
| Professional group (Medical staff) | 0.18 | 0.88 | 0.051 |
| Professional group (Others) | 0.52 | 0.53 | 0.222 |
| Transmission type (Possibly nosocomial) | 1.29 | 0.58 | 0.658 |
| Transmission type (Definitely nosocomial) | 3.05 | 0.57 | **0.049** |
| Work on the COVID ward (yes) | 1.24 | 0.55 | 0.700 |
| Initial Ct value < 30 | 3.36 | 0.53 | **0.022** |
| Rhinitis within the first 10 days | 1.06 | 0.57 | 0.922 |
| Concentration disorders within the first 10 days | 2.28 | 0.64 | 0.197 |
| Sore throat within the first 10 days | 0.33 | 0.56 | 0.051 |
| Fever within the first 10 days | 2.81 | 0.59 | 0.079 |
| Cough within the first 10 days | 1.38 | 0.54 | 0.556 |
| Breathlessness within the first 10 days | 7.89 | 0.78 | **0.008** |
| Shortness of breath within the first 10 days | 1.09 | 0.55 | 0.878 |
| Limb pain within the first 10 days | 1.16 | 0.63 | 0.814 |
| Unspecific pain within the first 10 days | 2.17 | 0.74 | 0.297 |
| Sleep disorder within the first 10 days | 1.69 | 0.72 | 0.467 |
| Brain fog within the first 10 days | 0.70 | 0.61 | 0.554 |
| Anxieties within the first 10 days | 2.84 | 0.90 | 0.245 |
| Mood changes within the first 10 days | 0.78 | 1.37 | 0.856 |
| Anosmia or Ageusia within the first 10 days | 3.01 | 0.53 | **0.037** |
| Heaviness of limbs or Weakness in the limbs within the first 10 days | 0.63 | 0.68 | 0.488 |
| Fatigue or Headache within the first 10 days | 0.20 | 0.65 | **0.014** |

Note. SE; standard error.
Model results for odds of illness duration over 90 days. Only hospital employees with available initial Ct value were included (N=150). For nominal scaled variables, odds ratio refers to the reference characteristic (male gender, nursing staff, work on the non-COVID ward, community acquired, initial Ct value >30). Odds ratios were calculated for the presence of the listed symptoms within the first ten days after initial SARS-CoV-2 detection compared to the absence of the respective symptoms. Symptoms that occurred rarely were not considered. If two symptoms correlate strongly with each other, they are combined and the combined symptom is called observed as soon as one of the symptoms occurs. The accuracy data of logistic regression model were as followed: accuracy 0.73, cross-validation accuracy 0.61, sensitivity 0.79, cross-validation sensitivity 0.53, specificity 0.64, cross-validation specificity 0.50.

Logistic regression was used to identify factors that increased the probability of illness duration lasting more than 90 days. The occurrence of breathlessness within the first ten days increased the probability of a prolonged illness duration over 90 days by a factor of OR = 7.89 (p = 0.008), the presence of anosmia or ageusia increased the probability by a factor of OR = 3.01 (p = 0.037). Initial Ct value < 30 (OR = 3.36; p = 0.022) and a definitely nosocomial SARS-CoV-2 transmission (OR = 3.05; p = 0.049) also showed a statistically significant correlation with increased odds of illness duration over 90 days.

## Discussion

In our cohort of SARS-CoV-2 positive tested and almost entirely still unvaccinated hospital employees during the first three waves (wild-type and alpha variant of SARS-CoV-2), a standardized interview revealed a high proportion of hospital employees with long lasting symptoms. Only twelve percent reported no symptoms at all, whilst 73% had symptoms longer than 28 days and 47% longer than 90 days. This goes in line with the work of Carvalho-Schneider et al., based on a prospective follow-up of 150 adults with noncritical COVID-19, that 68% of adults reported at least one lasting symptom at day 30 after symptom onset.^16^ Symptoms could be grouped in those lasting shorter, mid and longer times. Short-lasting symptoms were fever, rhinitis, sore throat and limb pain. Long-lasting symptoms were alopecia, concentration and memory disorder, palpitations, breathlessness, shortness of breath and sleep disorder. Symptoms could be grouped into symptom complexes. Two previous studies about long-lasting symptoms after SARS-CoV-2 infections in patient cohorts, using self-reporting questionnaire information, both described dry cough, diarrhea, confusion and disorientation, pain, anosmia, ageusia, and fatigue as long-lasting symptoms.^7,9^

Comparison of different observational studies upon long lasting symptoms after non–severe SARS-CoV-2 infection revealed differences in the studied cohorts, the items of the interviews and the follow up. Whereas the first studies included patients who were asked for convalescent plasma donation or patients who were surveyed in the context of a quality management system,^7,9^ here we included all employees of the Cologne Municipal Hospitals, representing the employable population. Employees were not only tested for SARS-CoV-2 if symptoms were present, but also all direct contact persons were actively screened for SARS-CoV-2 for 14 days after the documented contact. Hence, we were able to also include HEs with an asymptomatic course. Interestingly, only twelve percent of all employees had no symptoms at all. A share of 28% of employees included in the study were proven direct contacts to SARS-CoV-2 positive cases within the hospital, defined as definite nosocomial, and in additional 20% of cases, direct contacts could not be excluded, defined as possible nosocomial. Possible and definite nosocomial cases could be seen as a surrogate for a screening cohort and one could assume that in the screening cohort the proportion of HEs without symptoms could be higher. Thus, we would certainly have detected real asymptomatic cases in just under one half of our cohort, if asymptomatic courses were frequent – which was not the case.

Compared to large register studies like Roessler et al. where disease diagnoses were recorded,^17^ in the present study we were able to capture symptoms that were not necessarily linked to a physician’s disease diagnosis. These symptoms may nevertheless be essential to understand the burden of post-COVID conditions.^6^

In comparison to the work of Augustin et al. with a prevalence of 35% for six months lasting symptoms,^7^ in our cohort the prevalence of long-lasting symptoms after three months was at 47% of hospital employees, hence, being comparable. Augustin et al. defined the occurrence of at least one long lasting symptom out of four (anosmia, ageusia, fatigue, shortness of breath) as post-COVID syndrome.^7^ In our survey we included symptom features of Augustin et al. and Jones et al.,^7,9^ and added those symptom features we got reported during our individual phone calls, which were brain fog, weakness and heaviness of limbs.

Just recently, a nationwide cross-sectional study in the Danish population with over 60.000 SARS-CoV-2 positive persons and a SARS-CoV-2 negative control group revealed significant risk differences for anosmia, fatigue/exhaustion, dyspnea and reduced strengths in legs/arms.^6^ Our data, which do not include control groups, are consistent with the findings of the Danish questionnaire study.

In addition, we asked our employees when each symptom started and how long it lasted. These data we used to build a network of symptoms evolving over time. The network revealed symptom complexes until 28 days after infection combining sore throat, cough, anosmia, ageusia and fatigue. The symptom network of symptoms over 28 days identified headache, anosmia, ageusia, fatigue, concentration and memory disorders as well as shortness of breath as the most common combined symptoms. Over time, the symptom complexes changed. This is demonstrated with a movie of the evolving network in the supplemental material. Describing the post-COVID syndrome obviously depends on the time after the infection and the combination of different symptoms. Any symptom may occur solitarily or in any combination, which should be considered in post-COVID definitions and for a structured work up of a patient after SARS-CoV-2 infection.

Different symptom complexes following SARS-CoV-2 infection may be linked to different pathologies.^18^ Thus, Zazhytska et al. reported that damaged cerebral cells with persistent viral loads in some individuals being possibly the cause for mental disorders, or a disruption of nuclear architecture may be a cause of anosmia.^19^ Etter et al. showed signs of neurodegeneration associated to autoimmunity and peripheral immune signatures as possible causes for concentration and memory disorders or other central neurological symptoms.^20^ Persistent infection or inflammation of heart cells may be the reason for palpitation and breathlessness.^21,22^ Furthermore, data of the electronic database of the US Department of Veterans Affairs indicated that individuals with COVID-19 had an increased risk for different cardiovascular diseases in comparison to controls.^23^

Our study may be limited by a lacking control group in our study design. Considering the question, if long-lasting symptoms were caused by SARS-CoV-2 infection or did occur by chance, post-COVID symptoms were compared between SARS-CoV-2 positive and negative groups. The Danish cross-sectional study by Vedel Sørensen et al. as well as the findings of Roessler et al. confirmed our data.^6,17^ Roessler et al. conducted a matched cohort study based on routine health insurance data of more than 45 percent of the German population with a COVID-19 cohort including nearly 160.000 individuals.^17^ They defined 96 potential post-COVID health outcomes that consisted of new-onset diagnoses which were documented by a physician or psychotherapist. Outcomes were combined into 13 organ-related diagnosis/symptom complexes. Roessler et al. figured a higher incidence of new-onset diagnosis of all diagnosis/symptom complexes compared to matched adult control groups.

As an additional limitation of our study, it must be mentioned that we only asked about subjectively perceived symptoms that were not necessarily confirmed by a physician. However, the work of Roessler et al. confirms our findings and shows that also even more severe conditions that were diagnosed by a trained physician occur more often in the COVID-19 cohort than in the control cohort.^17^

Recently, Cohen et al. detected an increase of 11% for SARS-CoV-2 infected patients over the age of 65 for other sequelae.^24^ In our questionnaire, we explicitly asked only about new onset symptoms. Nevertheless, it must be mentioned that we can not exclude that some employees may have overstated their reported symptoms.

The socioeconomic impact of long lasting symptoms after a SARS-CoV-2 infection was early described by the surrogate sick leave of working people in Sweden. Median leave time from work was 35 days and 9% had sick leaves longer than four months.^10^

For both medical and socioeconomic reasons, predictors for long lasting symptoms are essential. Predictors were first identified by the Swedish study investigating sick leaves, which were older age and sick leaves within the year prior to infection in a first rough analysis.^10^

We identified a significant association between the occurrence of ageusia, anosmia or breathlessness during the first ten days, first Ct value <30 as well as a definitely nosocomial SARS-CoV-2 transmission, and a prolonged illness duration over 90 days. Through the identification of risk factors for long-lasting COVID-19 course, individuals who are at increased risk can be selected and medically treated at an early stage.^5^ An early identification of risk factors is important because drug therapy for COVID-19, for example monoclonal antibody therapy or antiviral therapy with Nirmatrelvir/Ritonavir (Paxlovid®), for example, should be carried out within the first 5 days after beginning of COVID-19 symptoms.^25^

Kuodi et al. examined the effectiveness of COVID-19 vaccines against long-term COVID-19 symptoms and showed, that vaccination with at least two doses of COVID-19 vaccine reduced the odds for post-COVID symptoms substantially and even bringing it to the same baseline like individuals reporting no previous SARS-CoV-2 infection.^26^ Therefore, the importance of full vaccination against COVID-19 must be emphasized also with regard to the prevention of long-lasting COVID-19 symptoms.

Surprisingly, the nosocomial acquisition was a significant predictor for a long lasting course. We may speculate the acquisition in a nosocomial context as a surrogate for a higher infectious dose. This is consistent with our finding of the association between a lower initial Ct value and a prolonged course.

## Conclusion

In line with recent literature we found that long-lasting symptoms after acute COVID-19 occur frequently, even after initially mild courses. About half of the HEs developed symptoms lasting more than 90 days after almost entirely mild courses of COVID-19. Different symptom complexes that developed and changed over time could be shown. According to our data, long-lasting COVID-19 symptoms can be either single symptoms or symptom complexes. Low initial Ct value, definitely nosocomial SARS-CoV-2 transmission and the presence of breathlessness, anosmia or ageusia within the first 10 days were identified as predictive factors for long-term courses of COVID-19. Predictive factors at the onset of the infection could possibly be used for early treatment to prevent development of long-term symptoms after COVID-19 in future.

## Supporting information

Questionnaire on persistent COVID-19 symptoms

Co-occurrence network of symptoms movie

## Data Availability

All data produced in the present study are available upon reasonable request to the authors.

## Author contributions

The study was designed and initiated by: FM, GS.

The questionnaire was designed by: FM, VM, CW.

Collection of data including programming related to this: VM, CW, FM, RG, GS.

Data analysis was done by: SJ, SH.

The first draft was written by: RG, FM.

All authors have critically revised the manuscript.

All authors have approved the final version of the manuscript and agreed to be accountable for all aspects of the work.

## Acknowledgements

The authors would like to thank all hospital employees for taking part in the survey. We also thank all staff of the Occupational Health Service as well as the Institute of Hygiene, especially our infection control nurses Heyde Brauell, Daniela Cöllen, Ina Dombrowski, Eva Ellers, Margit Frantzen, Gudrun Gaksch, Regine Galante, Andreas Kirchler, Andreas Kreutzberger, Stefanie Maczewski, Bilgen Pehlivan and Stefanie Teves, for their great commitment during the whole period of the survey.

## References

1. RKI. Robert Koch-Institut: COVID-19 Dashboard. 2021. https://experience.arcgis.com/experience/478220a4c454480e823b17327b2bf1d4 (accessed 17.02.2022).

2. WHO Working Group on the Clinical Characterisation and Management of COVID-19 infection. A minimal common outcome measure set for COVID-19 clinical research. The Lancet infectious diseases 2020; 20(8): e192–e7.

3. Huang C, Huang L, Wang Y, et al. 6-month consequences of COVID-19 in patients discharged from hospital: a cohort study. Lancet 2021; 397(10270): 220–32.

4. Nasserie T, Hittle M, Goodman SN. Assessment of the Frequency and Variety of Persistent Symptoms Among Patients With COVID-19: A Systematic Review. JAMA network open 2021; 4(5): e2111417.

5. Groff D, Sun A, Ssentongo AE, et al. Short-term and Long-term Rates of Postacute Sequelae of SARS-CoV-2 Infection: A Systematic Review. JAMA network open 2021; 4(10): e2128568.

6. Vedel Sørensen AI, Spiliopoulos L, Bager P, et al. Post-acute symptoms, new onset diagnoses and health problems 6 to 12 months after SARS-CoV-2 infection: a nationwide questionnaire study in the adult Danish population. medRxiv 2022: 2022.02.27.22271328.

7. Augustin M, Schommers P, Stecher M, et al. Post-COVID syndrome in non-hospitalised patients with COVID-19: a longitudinal prospective cohort study. The Lancet regional health Europe 2021; 6: 100122.

8. World Health Organization. A clinical case definition of post COVID-19 condition by a Delphi consensus. 2021. https://www.who.int/publications/i/item/WHO-2019-nCoV-Post_COVID-19_condition-Clinical_case_definition-2021.1.

9. Jones R, Davis A, Stanley B, et al. Risk Predictors and Symptom Features of Long COVID Within a Broad Primary Care Patient Population Including Both Tested and Untested Patients. Pragmatic and observational research 2021; 12: 93–104.

10. Westerlind E, Palstam A, Sunnerhagen KS, Persson HC. Patterns and predictors of sick leave after Covid-19 and long Covid in a national Swedish cohort. BMC public health 2021; 21(1): 1023.

11. R Core Team. R: A Language and Environment for Statistical Computing. 2021. https://www.R-project.org/.

12. Epskamp S, Cramer AOJ, Waldorp LJ, Schmittmann VD, Borsboom D. qgraph: Network Visualizations of Relationships in Psychometric Data. J Stat Soft 2012; 4: 1–18.

13. Therneau TM. A Package for Survival Analysis in R. 2021. https://CRAN.R-project.org/package=survival.

14. Cox DR. Regression Models and Life Tables. J R Stat Soc 1972; 34: 187–202.

15. Agresti A. Categorical Data Analysis. 2nd ed. New York: John Wiley & Sons; 2002.

16. Carvalho-Schneider C, Laurent E, Lemaignen A, et al. Follow-up of adults with noncritical COVID-19 two months after symptom onset. Clinical microbiology and infection : the official publication of the European Society of Clinical Microbiology and Infectious Diseases 2020; 27(2): 258–63.

17. Roessler M, Tesch F, Batram M, et al. Post COVID-19 in children, adolescents, and adults: results of a matched cohort study including more than 150,000 individuals with COVID-19. medRxiv 2021: 2021.10.21.21265133.

18. Mehandru S, Merad M. Pathological sequelae of long-haul COVID. Nature immunology 2022; 23(2): 194–202.

19. Zazhytska M, Kodra A, Hoagland DA, et al. Non-cell-autonomous disruption of nuclear architecture as a potential cause of COVID-19-induced anosmia. Cell 2022.

20. Etter MM, A. Martins T, Kulsvehagen L, et al. Severe Neuro-COVID is associated with peripheral immune signatures, autoimmunity and signs of neurodegeneration: a prospective cross-sectional study. medRxiv 2022: 2022.02.18.22271039.

21. Raveendran AV, Jayadevan R, Sashidharan S. Long COVID: An overview. Diabetes & metabolic syndrome 2021; 15(3): 869–75.

22. Yong SJ. Long COVID or post-COVID-19 syndrome: putative pathophysiology, risk factors, and treatments. Infectious diseases 2021; 53(10): 737–54.

23. Xie Y, Xu E, Bowe B, Al-Aly Z. Long-term cardiovascular outcomes of COVID-19. Nature medicine 2022.

24. Cohen K, Ren S, Heath K, et al. Risk of persistent and new clinical sequelae among adults aged 65 years and older during the post-acute phase of SARS-CoV-2 infection: retrospective cohort study. BMJ 2022; 376: e068414.

25. Robert Koch-Institut. Medikamentöse Therapie bei COVID-19 mit Bewertung durch die Fachgruppe COVRIIN beim Robert Koch-Institut. 2022. https://www.rki.de/DE/Content/InfAZ/N/Neuartiges_Coronavirus/COVRIIN_Dok/Therapieuebersicht.pdf?blob=publicationFile (accessed 10.03.2022).

26. Kuodi P, Gorelik Y, Zayyad H, et al. Association between vaccination status and reported incidence of post-acute COVID-19 symptoms in Israel: a cross-sectional study of patients tested between March 2020 and November 2021. medRxiv 2022: 2022.01.05.22268800.

