## Supplementary material for "Long-term symptoms after SARS-CoV-2 infection in a cohort of hospital employees: duration and predictive factors": Questionnaire on persistent COVID-19 symptoms

**Supplemental Material**

**Questionnaire For Hospital Employees On Persistent COVID-19 symptoms**

Patient/Hospital Employee (last name, first name, date of birth):________________________________________________________________

Date of survey:___________Who has surveyed?_____________

Has the survey been agreed to? Yes/No                                        Was the survey broken off? Yes/No

How was the course of your SARS-CoV-2 infection?

________________________________________________________________________________________(free text, use back page if necessary)

At what point did you feel fully fit again?  _____________________(date)

What do/did you suffer from (beyond preexisting symptoms prior your SARS-CoV-2 infection)?

1. Fatigue    no/yes: from__________until__________manifestation:_____________________________________________________
2. Sleep disorder    no/yes: from__________until__________manifestation:_____________________________________________________
3. Anosmia    no/yes: from__________until__________manifestation:_____________________________________________________
4. Ageusia    no/yes: from__________until__________manifestation:_____________________________________________________
5. Headache    no/yes: from__________until__________manifestation:_____________________________________________________
6. Dysesthesia    no/yes: from__________until__________manifestation:_____________________________________________________
7. Brain fog    no/yes: from__________until__________manifestation:_____________________________________________________
8. Concentration disorders no/yes: from__________until__________manifestation:____________________________________________________
9. Memory disorder    no/yes: from__________until__________manifestation:_____________________________________________________
10. Anxieties    no/yes: from__________until__________manifestation:_____________________________________________________
11. Mood changes    no/yes: from__________until__________manifestation:_____________________________________________________
12. Limb pain    no/yes: from__________until__________manifestation:_____________________________________________________

13. Heaviness of limbs no/yes: from____________until____________manifestation:________________________________________________

14. Unspecific pain no/yes: from____________until____________manifestation:________________________________________________

15. Weakness in the limbs no/yes: from____________until____________manifestation:________________________________________________

16. Sore throat no/yes: from____________until____________manifestation:________________________________________________

17. Fever no/yes: from____________until____________manifestation:________________________________________________

18. Alopecia no/yes: from____________until____________manifestation:________________________________________________

19. Rhinitis no/yes: from____________until____________manifestation:________________________________________________

20. Palpitations no/yes: from____________until____________manifestation:________________________________________________

21. Breathlessness no/yes: from____________until____________manifestation:________________________________________________

22. Shortness of breath no/yes: from____________until____________manifestation:_________________________ How many stairs can you walk? ______

23. Cough no/yes: from____________until____________manifestation:________________________________________________

24. Skin alterations no/yes: from____________until____________manifestation:__________________ What kind?_____________________________

25. Others? no/yes: from____________until____________manifestation:__________________ What kind?_____________________________

*Institute of Hygiene/Occupational Health Service. Questionnaire developed based on previously published studies*[1-3]

Literature sources:


1. Augustin M, Schommers P, Stecher M, et al. Post-COVID syndrome in non-hospitalised patients with COVID-19: a longitudinal prospective cohort study. *The Lancet regional health Europe* 2021; **6**: 100122.
2. Jones R, Davis A, Stanley B, et al. Risk Predictors and Symptom Features of Long COVID Within a Broad Primary Care Patient Population Including Both Tested and Untested Patients. *Pragmatic and observational research* 2021; **12**: 93-104.
3. Westerlind E, Palstam A, Sunnerhagen KS, Persson HC. Patterns and predictors of sick leave after Covid-19 and long Covid in a national Swedish cohort. *BMC public health* 2021; **21**(1): 1023.
