## Supplementary figures and images for "Long-term symptoms after SARS-CoV-2 infection in a cohort of hospital employees: duration and predictive factors"

### Co-occurrence network of symptoms movie

Co−occurence network of symptoms: Symptom days < 7

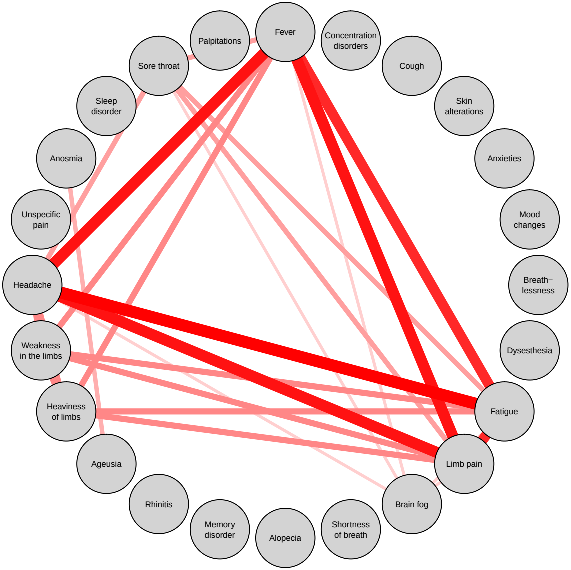


Play Pause
